# Malaria burden and associated risk factors in an area of pyrethroid-resistant vectors in southern Benin

**DOI:** 10.1101/2022.03.10.22272217

**Authors:** Manfred Accrombessi, Martin C. Akogbeto, Edouard Dangbenon, Hilaire Akpovi, Arthur Sovi, Boulais Yovogan, Constantin Adoha, Landry Assongba, Aurore Ogouyemi-Hounto, Germain Gil Padonou, Charles Thickstun, Mark Rowland, Corine Ngufor, Natacha Protopopoff, Jackie Cook

**Affiliations:** Faculty of Infectious and Tropical Diseases, Disease Control Department, London School of Hygiene and Tropical Medicine, WC1E 7HT, London, UK; Centre de Recherche Entomologique de Cotonou (CREC), Cotonou, Benin; UER Parasitology Mycology, Health Science Faculty, Abomey-Calavi University; National Malaria Control Program, Ministry of Health, Cotonou, Benin; School of Epidemiology and Public Health, Faculty of Medicine, University of Ottawa, Ottawa, Canada; Medical Research Council (MRC) International Statistics and Epidemiology Group, London School of Hygiene and Tropical Medicine, WC1E 7HT, London, UK

**Keywords:** Epidemiology, malaria infection, pyrethroid-insecticide resistance, risk factors, Benin

## Abstract

Malaria remains the main cause of morbidity and mortality in Benin despite the scale-up of long-lasting insecticidal nets (LLINs), indoor residual spraying, and malaria case management. This study aimed to determine the malaria burden and its associated risk factors in a rural area of Benin characterized by high net coverage and pyrethroid-resistant mosquito vectors. A community-based cross-sectional survey was conducted in three districts in southern Benin. Approximately 4,320 randomly selected participants of all ages were tested for malaria using rapid diagnostic tests within 60 clusters. Risk factors for malaria infection were evaluated using mixed-effect logistic regression models. Despite high population net use (96%), malaria infection prevalence was 43.5% (cluster range: 15.1-72.7%). Children (58.7%) were more likely to be infected than adults (31.2%), with a higher malaria prevalence among older children (5-10 years: 69.1%; 10-15 years: 67.9%) compared to young children (<5 years: 42.1%); however, young children were more likely to be symptomatic. High household density, low socioeconomic status, young age (<15years), poor net conditions, and low net usage during the previous week were significantly associated with malaria infection. Malaria prevalence remains high in this area of intense pyrethroid resistance despite high net use. New classes of LLINs effective against resistant vectors are therefore crucial to further reduce malaria in this area.

## Introduction

During the past few decades, long-lasting insecticidal nets (LLINs) have significantly contributed to the decline in malaria morbidity and mortality across sub-Saharan Africa (SSA).^1^ However, the progress achieved so far has stalled in many countries, with some seeing an increased burden in the last few years.^2^ COVID-19-related disruptions in the provision of malaria prevention, diagnosis, and treatment appear to have contributed to an increase in the number of cases in 2020.^3^ In Benin, malaria remains a serious public health issue despite the scale-up of LLINs, indoor residual spraying, and malaria case management.^4,5^ According to the national health statistics, 2,719,608 malaria cases were recorded in 2019 representing 45.5% of the overall outpatient diagnoses with 3,509 deaths attributable to malaria.^6^ In 2018, more than 90% of households declared to have at least one insecticide-treated net, and 71% had slept under nets the night before the survey.^7^

The emergence of insecticide resistance in malaria vectors, particularly to pyrethroids, raises concerns about the continued effectiveness of pyrethroid-only LLINs.^8–10^ In Benin, several studies have highlighted that a high proportion of malaria vectors are resistant to the pyrethroids used on insecticide-treated nets, including permethrin, deltamethrin, and alpha-cypermethrin,^11–15^ resulting in low mosquito mortality rates.^16,17^ However, the precise impact of resistance on malaria vector control efficacy is not fully known.^18–20^ In addition to insecticide resistance, a recent review has suggested that the resurgence of malaria cases may also be due to reductions in net usage and sub-standard nets.^21^

Due to concerns about the potential failure of current control tools, new types of LLINs have been developed to sustain malaria infection reduction.^22,23^ In preparation for a trial testing these new classes of LLINs, this study assessed the malaria infection burden and associated risk factors in Southern Benin, a malaria-endemic setting with pyrethroid-resistant malaria vectors.

## Materials and methods

### Study area and design

A community-based cross-sectional survey was conducted between October and November 2019 in three districts (Covè, Zagnanado, and Ouinhi) located in Southern Benin. The survey was part of pre-intervention activities of a cluster randomized controlled trial designed to assess the impact of 2 novels dual-active ingredient LLINs (Royal Guard^®^ net combining alpha-cypermethrin and pyriproxyfen; and Interceptor^®^ G2 net incorporating alpha-cypermethrin and chlorfenapyr) for control of malaria transmitted by pyrethroid-resistant vectors, a part of the “New Nets Project”. The trial protocol has been fully described elsewhere.^24^

Malaria is highly endemic in the study area and malaria transmission occurs year-round. There are two rainy seasons, from April to July and from October to November. Malaria prevalence in the region was 36.5% in children under 5 years old according to the DHS in 2018.^7^ The main vector control consists of the distribution of pyrethroid-only LLINs through universal coverage campaigns every 3 years (the most recent conducted in 2017) and routine service delivery to pregnant women during antenatal care and to children under 5 years during extended programs on immunization. An entomological survey conducted concurrently with this study,^25^ showed that *Anopheles coluzzii* and *Anopheles gambiae s*.*s*. were the main malaria vectors and both were highly resistant to pyrethroids mediated by multiple resistance mechanisms mainly L1014F *kdr* mutation (>80%) and over-expressed mixed-function oxidases. The indoor and outdoor entomological inoculation rate (EIR) was 21.6 and 5.4 infected bites/person/month, respectively.^25^

### Study population

A census was carried out in June 2019 in the 123 villages of the study area. Houses were numbered, georeferenced, and information on the household’s residents (name, sex, age, number of sleeping areas and LLINs, relationship with the household head) were collected. Sixty clusters were identified with each cluster comprising of 1 village or a group of villages for an average of 200 households (approximately 1200 residents). All resident adults or children over 6 months of age willing to participate and providing consent (or guardian’s consent) were eligible for the study.

### Study procedures

To achieve adequate participant enrolment, enhanced community sensitization activities were conducted before and during the survey with the support of hamlet leaders and community health workers. For malaria infection prevalence assessment, 72 individuals per cluster were randomly selected (total of 4320 participants), stratified by age (<5 years, 5-9 years, 10-14 years, ≥15 years) from each of the 60 clusters.

The survey included two components; (i) a household survey and (ii) a clinical survey. During the household survey, a questionnaire was administered to obtain information on possible risk factors for malaria and sampled persons were tested for malaria using rapid diagnostic tests (Malaria Pf HRP2 Ag RDT, Carestart^®^).

#### Household survey

Age-stratified individuals were randomly selected using two-stage cluster sampling from the census list generated during the registration activity. A maximum of two members was selected per house. The household questionnaire included information on gender, age, educational status and occupation, socioeconomic status, vector control measures used (number of nets, type and condition of nets, duration of usage), previous malaria cases, and care-seeking behaviors. The physical conditions of nets was assessed visually and categorized as good, average, and poor condition based on consensus between two field staff. We also assessed the presence of potential breeding sites such as water taps, open gutters, semi-open gutters, wells, puddles, septic tanks, or rivers in the proximity of the household.

#### Malaria testing

Participants were tested with an RDT, regardless of symptoms. A temperature record was taken for all participants using an infra-red thermometer. Symptomatic malaria infection was defined by a positive RDT plus fever (temperature ≥37.5°C) or a history of fever in the last 48 hours. Treatment was provided free of charge if they were found positive. Any participants with severe malaria or any diagnosed ailments that could not be treated by study staff were referred to the nearest health facilities.

Hemoglobin levels were measured with a Haemocue device in children aged 5 years old and under. Anemia was defined as a hemoglobin concentration below 110 g/L. Severe, moderate, and mild anemia were respectively defined as hemoglobin levels less than 70 g/L, between 70-99 g/L and 100-109 g/L.^26^

LLIN coverage was assessed with the following indicators,^27,28^ i) *Household net ownership* defined as the proportion of households with at least one bed net; ii) *Household net access* which is the proportion of households with enough nets for every two family members (based on one net for every two members); iii) *Population net access* corresponding to the proportion of individuals with access to bed nets within the households (assuming that each bed net in a household can be used by two people), and iv) *Population net use* representing the proportion of individuals who reported sleeping under a bed net the previous night. Net usage was also assessed during the previous week (net used 0-4 nights i.e. few nights, 5-6 nights i.e. most nights, 7 nights i.e. all nights).

Mosquito collections were undertaken across all villages, between September and October 2019. In each village, four houses were selected for mosquito sampling using human landing catches (HLCs). To ensure the supervision of mosquito collectors, the first house was randomly selected from the study census list, while the other three were chosen within a radius of 15–20 meters around the first one. Four collectors were required per house (one indoor and one outdoor replaced by 2 others after 6 hours of collection). Details on entomological data collection as well as baseline characteristics have been fully detailed elsewhere.^25^

### Data management and statistical analysis

All data collected during the survey were recorded in electronic forms on smartphones installed with Open Data Kit (ODK) collect and analyzed with STATA version 16 (Stata Corp LP, College Station, TX, USA).

Descriptive statistics and 95% confidence intervals were used to summarize demographic data. Mixed effect logistic regression models were used to assess risk factors with clusters included as a random effect. Variables with *p*-values below 0.2 were included in multivariate analyses and were eliminated step by step using the backward selection procedure. Only variables whose *p*-values were less than 0.05 were retained in the final model. For variables with more than two categories, a *p*-value of the global test is given. Spatial similarities in malaria prevalence and EIR were examined visually using QGIS, with “natural breaks” to display the most variation in the data, given the histogram for each variable.

Risk factors assessed included characteristics at the household level (number of residents, head of the household gender and education, socio-economic status, ethnic group, net ownership, and access), and individual level (age, gender, net usage). Household socioeconomic status was created by using principal component analysis with the following variables included: type of lighting, access to water, type of roof, type of floor, type of toilet, household head level of education, household crowding, and ownership of assets (motorbike, television, bike, radio, sheep, bed, phone).

### Ethical Statement

The study protocol was approved by the Benin Ministry of Health ethics committee (Reference N°6/30/MS/DC/SGM/DRFMT/CNERS/SA) and by the institutional ethics review board of the London School of Hygiene and Tropical Medicine (N°16237). All participation was voluntary. Written informed consent was obtained from an adult guardian in the household or given by the participant if over 18 years. Assent was sought for children over 10 years.

## Results

A total of 216,138 inhabitants in 54,143 households were identified as part of the census. 4,441 randomly selected participants from 3,027 households were included in the baseline survey.

### Sociodemographic characteristics of the study population

The mean number of people living in households was 5.3 (Standard deviation [SD]±2.6) inhabitants. The number of sleeping spaces in the household was on average 2.7 (SD±1.3). The majority of heads of household had no formal education (71.4%) and the main ethnic group was “Fon” (77.2%). Mosquito breeding sites were observed close to 76.9% of households. Females represented 50.8% of the participants; children under 5 years old, 5-10 years, 10-15 years, and above to 15 years accounted for 16.5%, 14.4%, 13.6%, and 55.4%, respectively. Demographic, socioeconomic, and household characteristics of study participants are detailed in Table 1.

**Table 1.** Baseline characteristics of the study population

| Characteristics |  | Proportion or mean |
| --- | --- | --- |
| **Cluster-level** |  |  |
| Number of clusters |  | 60 |
| Number of households per cluster | Mean (range) | 904 (82-5403) |
| Proportion of children under 10 years per cluster, % | Mean (range) | 30.9 (25-35.2) |
| Cluster with high socioeconomic status, % (n) | Yes | 62.7 (37) |
| **Household-level** |  |  |
| Number of households enrolled |  | 3,027 |
| People living in the household | Mean (± SD) | 5.3 (± 2.6) |
| Number of sleeping spaces | Mean (± SD) | 2.7 (± 1.3) |
| Head of household gender, % (n) | Male | 83.4 (2,525) |
|  | Female | 16.6 (502) |
| Head of household education level, % (n) | No education | 71.4 (2,160) |
|  | Primary education | 17.2 (521) |
|  | Lower secondary education | 7.3 (220) |
|  | Higher secondary education | 2.7 (81) |
|  | College/University | 1.4 (45) |
| Ethnic group, % (n) | Fon | 77.2 (2,336) |
|  | Other* | 22.8 (691) |
| Presence of potential breeding sites, % (n) | Yes | 76.9 (2,209) |
| **Individual-level** |  |  |
| Number of participants enrolled |  | 4,441 |
| Participant gender, % (n) | Male | 49.1 (2,182) |
|  | Female | 50.8 (2,259) |
| Age, years % (n) | $\leq 5$ | 16.5 (735) |
|  | 5-10 | 14.4 (638) |
|  | 10-15 | 13.6 (606) |
|  | > 15 | 55.4 (2,462) |
| Education level, % (n) | Not in the age of education | 16.5 (735) |
|  | Not educated | 52.5 (2,333) |
|  | Educated | 30.9 (1,373) |

Abbreviations: SD, standard deviation
\*Other ethnic groups: Mahi, Holli, Yoruba, Goun

### Household LLIN characteristics

Table 2 presents the characteristics of bed nets recorded in the households. The number of bed nets in the household was on average 2.5 (SD±1.3). Most of the nets found were insecticide-treated nets (96.5%). The majority of the bed nets were provided during the previous national campaign of 2017 (91%) with others supplied through public health routine activities (8%, from antenatal care visits and expanded program immunization). PermaNet^®^ 2.0 was the most common LLIN type observed in the households (64.3%). Most bed nets had been used for more than two years (92.7%). The proportion of LLIN in poor condition was 37.9%. Household net ownership, household net access, population net access, and population net use were 95.8%, 55.9%, 79.5%, 95.8%, respectively. The majority of the participants (88.4%) declared to have slept under an LLIN every night during the previous week. Children under 5 years old had slept more often under a net every night the previous week in comparison to others (<5 years: 93.1%; 5-10 years: 88%; 10-15 years: 84.9%; >15 years: 87.3%).

**Table 2.** Characteristics of long-lasting insecticidal nets presented in the household

| Characteristics |  | Proportion or mean |
| --- | --- | --- |
| **Household-level** |  |  |
| Number of households enrolled |  | 3,027 |
| Household net ownership, % (n) | Yes | 95.8 (2,902) |
| Household net access, % (n) | Yes | 55.9 (1,691) |
| Population net access, % (95% CI) | Yes | 79.5 (78.9-80.1) |
| Number of nets (any type) in the household | Mean (± SD) | 2.5 (± 1.3) |
| Number of LLIN in the household | Mean (± SD) | 2.3 (± 1.3) |
| **Individual-level** |  |  |
| Number of participants enrolled |  | 4,441 |
| Number of participants enrolled with a net |  | 4,088 |
| Number of participants enrolled with an LLIN |  | 3,949 |
| Origin of LLIN, % (n) | National campaign (2017) | 91.0 (3,595) |
|  | Health care facility (ANC) | 6.4 (251) |
|  | Health care facility (EPI) | 1.6 (65) |
|  | Public market | 0.4 (16) |
|  | Other | 0.6 (22) |
| Proportion of LLIN observed, % (n) | Yes | 99.0 (3,910) |
| Type of LLIN, % (n) | Permanet 2.0 | 64.3 (2,489) |
|  | Dawa plus | 9.6 (374) |
|  | Olyset Net | 7.0 (271) |
|  | Duranet | 1.4 (53) |
|  | Yorkool | 0.5 (21) |
|  | Other* | 5.7 (221) |
|  | Unknown | 11.4 (443) |
| Duration usage of LLIN, % (n) | ≤1 year | 7.2 (286) |
|  | 2 years | 64.3 (2,540) |
|  | 3 years | 21.1 (833) |
|  | >3 years | 7.3 (290) |
| LLIN conditions, % (n) | Good | 18.9 (732) |
|  | Average | 43.2 (1,671) |
|  | Poor | 37.9 (1,468) |
| Population LLIN use, % (n) | Yes | 95.8 (3,782) |
| Net usage the week before the visit, % (n) | All nights | 88.4 (3,490) |
|  | Most of the nights | 8.6 (338) |
|  | Few nights | 3.1 (121) |

Abbreviations: SD, Standard deviation; LLIN, Long-lasting insecticidal net; ANC, Antenatal care visit; EPI, Expanded program Immunization
\* Most important other types of bed nets observed were impregnated by deltamethrin (88.2%, 195/221)

### Prevalence of malaria and analysis of potential risk factors

The prevalence of malaria infection was 43.5% (cluster range: 15.1-72.7%) (Table 3). Children less than 5 years old (malaria prevalence 42.1%), aged between 5-10 years (69.1%), and 10-15 years (67.9%) were most likely to be infected with malaria in comparison to adult participants (31.2%). However, among infected participants, the proportion of symptomatic cases was higher among children under 5 years (33.3%) in comparison to older children (5-10 years: 28.9%; 10-15 years: 23.7%) and adults (19.2%). The proportion of people presenting a fever or history of fever in the last 48 hours was 18.7% (829/4,441) of which 57.2% were malaria positive. Among children under 5 years old, anemia was present in 75.9% (cluster range: 33.3% to 100%).

**Table 3.** Malaria indicators in the study population

| Characteristics |  | Proportion<br>% (n) |
| --- | --- | --- |
| Number of participants enrolled, n |  | 4,441 |
| History of fever in the past 48 hours | Yes | 12.6 (562) |
| Fever* at the time of visit | Yes | 8.0 (356) |
| Hemoglobin test performed among children under 5 years | Yes | 99.5 (731) |
| Anemia<sup>\$</sup> among children under 5 years old | Yes | 75.9 (555) |
| Malaria rapid diagnostic test performed | Yes | 99.7 (4,428) |
| Malaria infection | Yes | 43.5 (1,924) |
| Malaria infection by age category | ≤ 5 years | 42.1 (309) |
|  | 5-10 years | 69.1 (440) |
|  | 10-15 years | 67.9 (410) |
|  | > 15 years | 31.2 (765) |
| Proportion of symptomatic cases<sup>#</sup> among infected per age | ≤ 5 years | 33.3 (103/309) |
|  | 5-10 years | 28.9 (127/440) |
|  | 10-15 years | 23.7 (97/410) |
|  | > 15 years | 19.2 (147/765) |
| Proportion of malaria among participants with fever or history of fever in the past 48 hours | Yes | 57.3 (474) |

\* Fever: infra-red frontal temperature $\geq 37.5^{\circ}\text{C}$
\$ Hemoglobin (Hb) $<110\text{ g/L}$

Household population density, socioeconomic status, gender, ethnic group, age, households with enough LLIN, net use the previous week, and net usage duration were factors associated with malaria infection (Table 4). After adjusting for confounding factors, the risk of malaria infection was significantly higher with an increasing number of people living in the house (adjusted odds ratio [aOR] 1.32, 95% CI 1.05-1.69 for households having between 3-7 members; and aOR 1.54, 95% CI 1.16-2.04 for households having more than 7 members in comparison to households with less than 3 members). People living in households with average (aOR1.43, 95%CI 1.19-1.69) and low (aOR 1.44, 95%CI 1.16-1.79) socioeconomic levels were more at risk of malaria infection than those living in households with high socioeconomic status. The age groups at highest risk were children under 5 years (aOR 1.66, 95% CI 1.38-1.99), between 5-10 years (aOR 5.31, 95% CI 4.33-6.53) and 10-15 years (aOR 4.88, 95% CI 3.96-6.03) in comparison to participants over 15 years. Participants who reported sleeping under nets a few nights (< 4 nights) the previous week were also more likely to be infected (aOR 1.95, 95% CI 1.29-2.94) in comparison to those who slept under nets every night. Also, household members using nets in poor condition were more likely to be infected (aOR 1.34, 95% CI 1.07-1.74) in comparison to those owning nets in good condition.

**Table 4.** Risk factors associated with malaria infection in an area of pyrethroid-resistant vectors in Benin

| Factors |  | Malaria positive % (n) | Univariate analysis |  | Multivariate analysis |  |
| --- | --- | --- | --- | --- | --- | --- |
|  |  |  | OR* (95% CI) | p-value | aOR** (95% CI) | p-value |
| Household-level |  |  |  |  |  |  |
| People living in the household | <3 | 29.2% (148) | 1 |  | 1 |  |
|  | 3-7 | 43.8% (1,382) | 1.82 (1.47-2.24) | <0.001 | 1.29 (1.01-1.65) | 0.03 |
|  | >7 | 51.6% (394) | 2.36 (1.84-3.02) |  | 1.47 (1.10-1.96) |  |
| Household socioeconomic status | High | 36.7% (542) | 1 |  | 1 |  |
|  | Average | 46.8% (685) | 1.32 (1.12-1.57) | 0.002 | 1.40 (1.12-1.75) | <0.001 |
|  | Low | 46.8% (697) | 1.33 (1.10-1.61) |  | 1.41 (1.17-1.72) |  |
| Household net access | No | 40.4% (989) | 1 |  |  |  |
|  | Yes | 47.2% (935) | 1.23 (1.08-1.39) | 0.002 |  |  |
| Individual-level |  |  |  |  |  |  |
| Gender | Female | 41.2% (928) | 1 |  |  |  |
|  | Male | 45.7% (996) | 1.18 (1.05-1.34) | 0.007 |  |  |
| Ethnic group | Other | 39.4% (408) | 1 |  |  |  |
|  | Fon | 44.7% (1,516) | 1.17 (0.95-1.45) | 0.13 |  |  |
| Participant age | > 15 years | 31.2% (765) | 1 |  | 1 |  |
|  | 10-15 years | 67.9% (410) | 5.15 (4.23-6.28) |  | 4.96 (4.01-6.13) |  |
|  | 5-10 years | 69.1% (440) | 5.43 (4.47-6.61) | <0.001 | 5.52 (4.48-6.81) | <0.001 |
|  | ≤ 5 years | 42.1% (309) | 1.66 (1.39-1.98) |  | 1.68 (1.39-2.03) |  |
| Duration usage of LLIN | ≤1 year | 37.4% (116) | 1 |  |  |  |
|  | 2 years | 43.7% (1,139) | 1.41 (1.07-1.84) |  |  |  |
|  | 3 years | 47.2% (408) | 1.58 (1.17-2.13) | 0.01 |  |  |
|  | >3 years | 37.0% (110) | 1.15 (0.78-1.70) |  |  |  |
| Net usage the previous week | All nights | 43.2% (1,552) | 1 |  | 1 |  |
|  | Most of nights | 43.3% (154) | 1.18 (0.91-1.54) | 0.04 | 1.01 (0.76-1.36) | 0.02 |
|  | Few nights | 51.5% (67) | 1.60 (1.09-2.35) |  | 1.83 (1.19-2.83) |  |
| Net conditions | Good | 38.4% (289) | 1 |  | 1 |  |
|  | Average | 42.9% (732) | 1.23 (0.99-1.53) | 0.002 | 1.23 (0.97-1.55) | 0.04 |
|  | Poor | 46.1% (705) | 1.48 (1.17-1.85) |  | 1.37 (1.07-1.74) |  |

Abbreviations: OR, odds ratio; aOR, adjusted odds ratio; LLIN, long-lasting insecticidal nets
\*Adjusted for within-cluster effect. \*\*Adjusted for significant variables associated with malaria infection and within-cluster effect

We observed a similar spatial distribution of malaria vector density and the prevalence of infection within clusters (Figure 1), with both indicators being high in the north of the study area. The average indoor monthly EIR in *An. gambiae* s.l. in clusters with malaria prevalence ≥50% was higher in comparison to clusters with malaria prevalence <50% (29.6 vs 13.6 bites/person/month).

**Figure 1.**
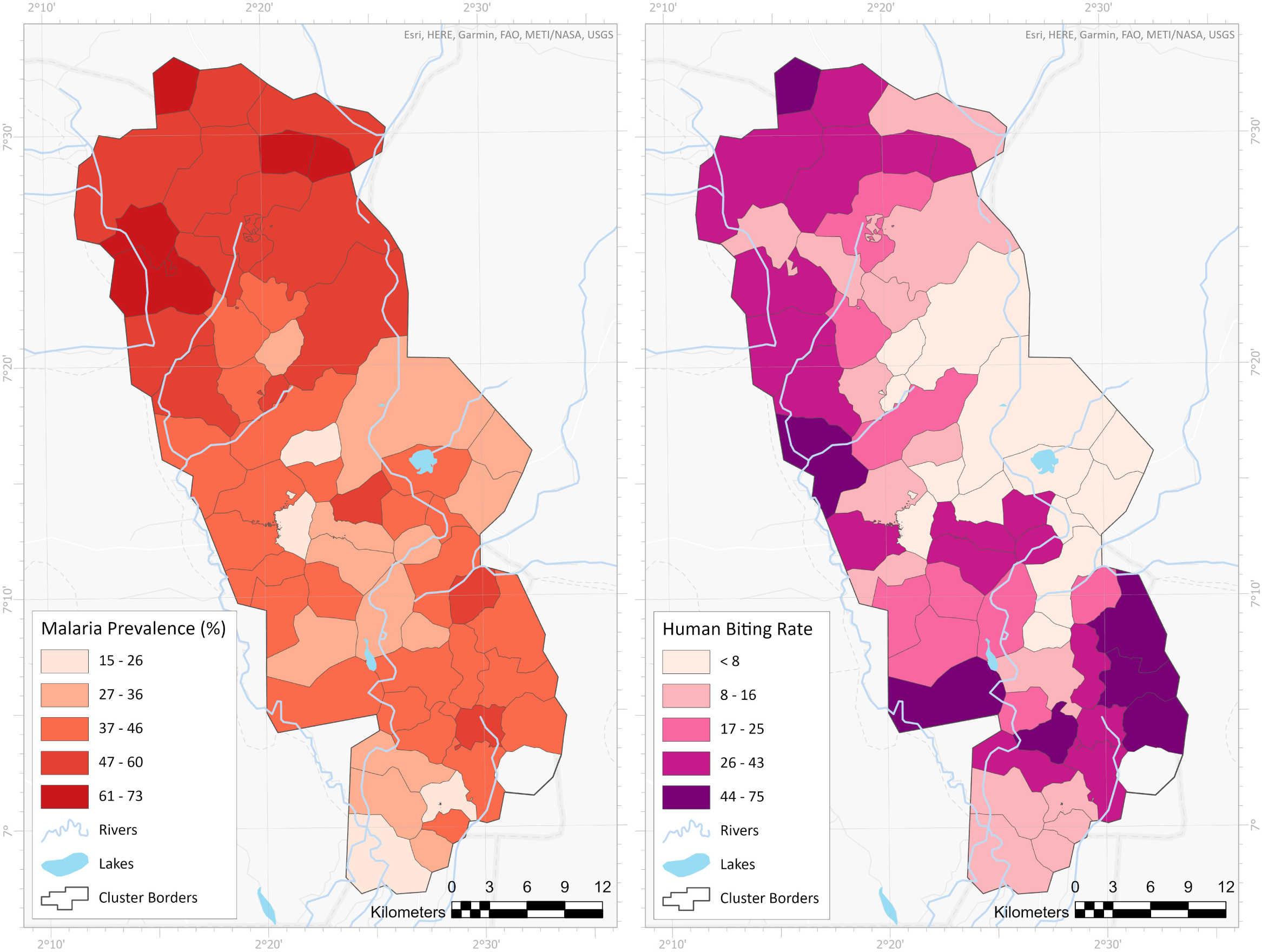
Comparison map of malaria prevalence and vector density within the 60 study clusters

## Discussion

This study examined risk factors for malaria infection in an area of intense transmission and the presence of pyrethroid-resistant vector mosquitoes.^15,16^ There is limited cross-sectional data available for this part of Benin and the current study represents a detailed analysis of the epidemiological picture, and compares it to the entomological outcomes reported in the same area.

Our data showed a high prevalence of malaria infection (43.5%) despite the high household net ownership and population net use. These findings are consistent with previous studies conducted in Benin.^29–31^Despite there being several years since the last universal net campaign, net ownership, access, and use remained very high. The concerningly high prevalence, despite high net use, may be partially explained by the high level of pyrethroid resistance in the local malaria vectors.^21,28^ Indeed, entomological surveys conducted simultaneously to this study have confirmed the scale of pyrethroid resistance with mosquitoes mortality rates at 24h, 48h, and 72h post-exposure lower than 40% for alphacypermethrin and permethrin insecticides and a frequency of the L1014F *kdr* mutation over 80%.^25^ Several studies have also highlighted the increasing role of pyrethroid-resistant *Anopheles* vectors in malaria transmission.^9,12,14,32^ However, this observation might be also related to the age and condition of the nets.^33^ In fact, we observed in this study that individuals using nets in poor condition were 1.37-fold more likely to be infected than those sleeping under nets in good condition.

Children under 15 years were most likely to be infected with malaria, with children aged between 5 and 15 years at higher risk than those under 5 years. Traditionally, children aged under 5 years have been thought to be at the highest risk of malaria infection. Changes in infection burden by age group have also been reported elsewhere in SSA.^34–36^ This has important implications for targeting interventions and surveillance in SSA. Several studies have previously suggested that reduced transmission might lead to increased susceptibility to malaria infection among older children due to lower acquired immunity.^37–39^ However, good malaria case management in children under 5 years old might also partially explain the shift in infection burden. Despite infection being higher in older children, the most severe health consequences continue to mainly affect the youngest children.^40,41^ Indeed, 33.3% of the youngest children (<5 years) tested for malaria had fever whereas 28.9%, 23.7% and 19.2% of infected children between 5-10 years, 10-15 years, and adults (>15 years) presented malaria related-morbidity, respectively. Furthermore, even though more asymptomatic, older children and adults are important drivers of malaria infection as they might not seek treatment and remain a reservoir for infection, undermining elimination efforts.^42–44^ They should, therefore, be considered for malaria public health interventions.

Socioeconomic status was also a factor significantly associated with malaria infection. Indeed, a recent systematic review has shown that an improved socioeconomic status would reduce dramatically the burden of malaria in SSA.^45^ Moreover, participants not sleeping under nets usually were more likely to be infected. This is in line with other studies highlighting the importance of population net usage despite the access.^21,46^

We observed a similar spatial distribution of malaria infection and malaria vector density and entomological inoculation rate over the clusters, highlighting the usefulness of both indicators.^47,48^

There is little information in Benin regarding the effect of increasing LLIN use and increasing insecticide resistance on malaria vector control and impact on malaria infection.^19,20,49^ These baseline results suggested that pyrethroid LLIN provide, at best, only partial protection against malaria and the importance of trialing novel LLIN technologies and approaches.^20,50^ The development and impact of insecticide resistance on malaria outcomes are key indicators to monitor. ^17,20,25,51,52^

LLINs remain a cornerstone of malaria control in Africa; however, the progress achieved so far is seriously threatened by pyrethroid-resistant malaria vectors. The increasing insecticide resistance in malaria vectors could have dramatic implications for malaria control and requires urgent remedial actions.^9^ It is, therefore, crucial to assess new classes of LLINs effective against resistant vectors to sustaining malaria reduction. Piperonyl butoxide-treated insecticidal nets have recently proven their effectiveness in malaria reduction in East Africa, though evidence of their epidemiological impact in West Africa is lacking.^53,54^ Other such novel generations of ITNs (pyrethroid-pyriproxyfen and pyrethroid-chlorfenapyr ITNs) are currently under evaluation in communities in Benin as part of the “New Nets Project which aims to accelerate the impact of these new technologies across sub-Saharan Africa.^22,23^

## Data Availability

All data produced in the present work are contained in the manuscript

## Declarations

## Acknowledgments

The authors gratefully acknowledge the study participants. We also thank the health community workers, local medical staff and community leaders of the Zou region for supporting the study and for agreeing to collaborate with the study investigators. Electronic data solutions were provided by LSHTM Open Research Kits (odk.lshtm.ac.uk). More detail on the New Nets Project is available at: https://www.ivcc.com/market-access/new-nets-project/.

## Authors’ contribution

MA, MCA, NP, and JC conceived and designed the study. MA, ED, HA, LA, AOH, and MCA implemented the study and supervised the data collections. MA, NP, and JC analyzed the data. CT created the study maps. MA, MCA, NP, and JC interpreted the data. MA wrote the initial draft of the manuscript which was firstly revised by NP, and JC. MA, MCA, ED, HA, AS, BY, CA, AOH, GGP, CT, CN, MR, NP, and JC critically revised the manuscript for important content. All authors read and approved the final manuscript.

## Authors’ addresses

*Faculty of Infectious and Tropical Diseases, Disease Control Department, London School of Hygiene and Tropical Medicine, WC1E 7HT, London, UK*

Manfred Accrombessi, Arthur Sovi, Corine Ngufor, Mark Rowland, Natacha Protopopoff.

*Centre de Recherche Entomologique de Cotonou (CREC), Cotonou, Benin*

Martin C. Akogbeto, Edouard Dangbenon, Hilaire Akpovi, Boulais Yovogan, Constantin Adoha, Landry Assongba, Germain Gil Padonou.

*National Malaria Control Program, Ministry of Health, Cotonou, Benin*

Aurore Ogouyemi-Hounto

*School of Epidemiology and Public Health, Faculty of Medicine, University of Ottawa, Ottawa, Canada*

Charles Thickstun

*Medical Research Council (MRC) International Statistics and Epidemiology Group, London School of Hygiene and Tropical Medicine, WC1E 7HT, London, UK*

Jackie Cook

## References

1. Bhatt S, Weiss DJ, Cameron E, Bisanzio D, Mappin B, Dalrymple U, Battle K, Moyes CL, Henry A, Eckhoff PA, Wenger EA, Briët O, Penny MA, Smith TA, Bennett A, et al., 2015. The effect of malaria control on Plasmodium falciparum in Africa between 2000 and 2015. Nature 526: 207–211

2. World Health Organization., 2020. World malaria report 2020: 20 tears of global progress and challenges. Geneva: World Health Organization; 2019. License: CC BY-NC-SA 3.0 IGO: 299

3. World Health Organization, 2021. World malaria report 2021. Geneva: World Health Organization, 2021 - License: CC BY-NC-SA 3.0 IGO. : 322

4. Akogbéto MC, Dagnon F, Aïkpon R, Ossé R, Salako AS, Ahogni I, Akinro B, Sominahouin A, Sidick A, Tokponnon F, Padonou GG., 2020. Lessons learned, challenges and outlooks for decision-making after a decade of experience monitoring the impact of indoor residual spraying in Benin, West Africa. Malar J 19: 45

5. Kleinschmidt I, Mnzava AP, Kafy HT, Mbogo C, Bashir AI, Bigoga J, Adechoubou A, Raghavendra K, Knox TB, Malik EM, Nkuni ZJ, Bayoh N, Ochomo E, Fondjo E, Kouambeng C, et al., 2015. Design of a study to determine the impact of insecticide resistance on malaria vector control: a multi-country investigation. Malar J 14: 282

6. National System of Health Information and Management (SNIGS), 2020. National health statistics 2019

7. National Institute of Statistic and Economic Analysis (INSAE), 2019. Demographic Health Survey in Benin, 2017-2018 : Key indicators. Cotonou, Bénin et Rockville, Maryland, USA : INSAE et ICF

8. Chandre F, Darrier F, Manga L, Akogbeto M, Faye O, Mouchet J, Guillet P., 1999. Status of pyrethroid resistance in Anopheles gambiae sensu lato. Bull World Health Organ 77: 230–234

9. Ranson H, Lissenden N., 2016. Insecticide Resistance in African Anopheles Mosquitoes: A Worsening Situation that Needs Urgent Action to Maintain Malaria Control. Trends Parasitol 32: 187–196

10. Djogbénou L, Pasteur N, Bio-Bangana S, Baldet T, Irish SR, Akogbeto M, Weill M, Chandre F., 2010. Malaria vectors in the Republic of Benin: distribution of species and molecular forms of the Anopheles gambiae complex. Acta Trop 114: 116–122

11. Akogbéto M, Yakoubou S., 1999. [Resistance of malaria vectors to pyrethrins used for impregnating mosquito nets in Benin, West Africa]. Bull Soc Pathol Exot 1990 92: 123– 130

12. Yadouleton AW, Padonou G, Asidi A, Moiroux N, Bio-Banganna S, Corbel V, N’guessan R, Gbenou D, Yacoubou I, Gazard K, Akogbeto MC., 2010. Insecticide resistance status in Anopheles gambiae in southern Benin. Malar J 9: 83

13. Aïkpon R, Sèzonlin M, Ossè R, Akogbéto M., 2014. Evidence of multiple mechanisms providing carbamate and organophosphate resistance in field An. gambiae population from Atacora in Benin. Parasit Vectors 7: 568

14. Gnanguenon V, Agossa FR, Badirou K, Govoetchan R, Anagonou R, Oke-Agbo F, Azondekon R, AgbanrinYoussouf R, Attolou R, Tokponnon FT, Aïkpon R, Ossè R, Akogbeto MC., 2015. Malaria vectors resistance to insecticides in Benin: current trends and mechanisms involved. Parasit Vectors 8: 223

15. Ngufor C, N’Guessan R, Fagbohoun J, Subramaniam K, Odjo A, Fongnikin A, Akogbeto M, Weetman D, Rowland M., 2015. Insecticide resistance profile of Anopheles gambiae from a phase II field station in Cové, southern Benin: implications for the evaluation of novel vector control products. Malar J 14: 464

16. Sagbohan HW, Kpanou CD, Osse R, Dagnon F, Padonou GG, Sominahouin AA, Salako AS, Sidick A, Sewade W, Akinro B, Ahmed S, Impoinvil D, Agbangla C, Akogbeto M., 2021. Intensity and mechanisms of deltamethrin and permethrin resistance in Anopheles gambiae s.l. populations in southern Benin. Parasit Vectors 14: 202

17. Kpanou CD, Sagbohan HW, Dagnon F, Padonou GG, Ossè R, Salako AS, Sidick A, Sewadé W, Sominahouin A, Condo P, Ahmed SH, Impoinvil D, Akogbéto M., 2021. Characterization of resistance profile (intensity and mechanisms) of Anopheles gambiae in three communes of northern Benin, West Africa. Malar J 20: 328

18. Bradley J, Ogouyèmi-Hounto A, Cornélie S, Fassinou J, de Tove YSS, Adéothy AA, Tokponnon FT, Makoutode P, Adechoubou A, Legba T, Houansou T, Kinde-Gazard D, Akogbeto MC, Massougbodji A, Knox TB, et al., 2017. Insecticide-treated nets provide protection against malaria to children in an area of insecticide resistance in Southern Benin. Malar J 16: 225

19. Tokponnon FT, Sissinto Y, Ogouyémi AH, Adéothy AA, Adechoubou A, Houansou T, Oke M, Kinde-Gazard D, Massougbodji A, Akogbeto MC, Cornelie S, Corbel V, Knox TB, Mnzava AP, Donnelly MJ, et al., 2019. Implications of insecticide resistance for malaria vector control with long-lasting insecticidal nets: evidence from health facility data from Benin. Malar J 18: 37

20. Kleinschmidt I, Bradley J, Knox TB, Mnzava AP, Kafy HT, Mbogo C, Ismail BA, Bigoga JD, Adechoubou A, Raghavendra K, Cook J, Malik EM, Nkuni ZJ, Macdonald M, Bayoh N, et al., 2018. Implications of insecticide resistance for malaria vector control with long-lasting insecticidal nets: a WHO-coordinated, prospective, international, observational cohort study. Lancet Infect Dis 18: 640–649

21. Lindsay SW, Thomas MB, Kleinschmidt I., 2021. Threats to the effectiveness of insecticide-treated bednets for malaria control: thinking beyond insecticide resistance. Lancet Glob Health 9: e1325–e1331

22. N’Guessan R, Odjo A, Ngufor C, Malone D, Rowland M., 2016. A Chlorfenapyr Mixture Net Interceptor® G2 Shows High Efficacy and Wash Durability against Resistant Mosquitoes in West Africa. PloS One 11: e0165925

23. Ngufor C, Agbevo A, Fagbohoun J, Fongnikin A, Rowland M., 2020. Efficacy of Royal Guard, a new alpha-cypermethrin and pyriproxyfen treated mosquito net, against pyrethroid-resistant malaria vectors. Sci Rep 10: 12227

24. Accrombessi M, Cook J, Ngufor C, Sovi A, Dangbenon E, Yovogan B, Akpovi H, Hounto A, Thickstun C, Padonou GG, Tokponnon F, Messenger LA, Kleinschmidt I, Rowland M, Akogbeto MC, et al., 2021. Assessing the efficacy of two dual-active ingredients long-lasting insecticidal nets for the control of malaria transmitted by pyrethroid-resistant vectors in Benin: study protocol for a three-arm, single-blinded, parallel, cluster-randomized controlled trial. BMC Infect Dis 21: 194

25. Yovogan B, Sovi A, Padonou GG, Adoha CJ, Akinro B, Chitou S, Accrombessi M, Dangbénon E, Akpovi H, Messenger LA, Ossè R, Hounto AO, Cook J, Kleinschmidt I, Ngufor C, et al., 2021. Pre-intervention characteristics of the mosquito species in Benin in preparation for a randomized controlled trial assessing the efficacy of dual active-ingredient long-lasting insecticidal nets for controlling insecticide-resistant malaria vectors. PloS One 16: e0251742

26. McLean E, Cogswell M, Egli I, Wojdyla D, de Benoist B., 2009. Worldwide prevalence of anaemia, WHO Vitamin and Mineral Nutrition Information System, 1993-2005. Public Health Nutr 12: 444–454

27. Seyoum D, Speybroeck N, Duchateau L, Brandt P, Rosas-Aguirre A., 2017. Long-Lasting Insecticide Net Ownership, Access and Use in Southwest Ethiopia: A Community-Based Cross-Sectional Study. Int J Environ Res Public Health 14: E1312

28. Roll Back Malaria Monitoring & Evaluation Reference Group., 2018. Household survey indicators for malaria control

29. Damien GB, Djènontin A, Rogier C, Corbel V, Bangana SB, Chandre F, Akogbéto M, Kindé-Gazard D, Massougbodji A, Henry M-C., 2010. Malaria infection and disease in an area with pyrethroid-resistant vectors in southern Benin. Malar J 9: 380

30. Nahum A, Erhart A, Mayé A, Ahounou D, van Overmeir C, Menten J, van Loen H, Akogbeto M, Coosemans M, Massougbodji A, D’Alessandro U., 2010. Malaria Incidence and Prevalence Among Children Living in a Peri-Urban Area on the Coast of Benin, West Africa: A Longitudinal Study. Am J Trop Med Hyg 83: 465–473

31. Wang S-J, Lengeler C, Smith TA, Vounatsou P, Akogbeto M, Tanner M., 2006. Rapid Urban Malaria Appraisal (RUMA) IV: Epidemiology of urban malaria in Cotonou (Benin). Malar J 5: 45

32. Matowo NS, Martin J, Kulkarni MA, Mosha JF, Lukole E, Isaya G, Shirima B, Kaaya R, Moyes C, Hancock PA, Rowland M, Manjurano A, Mosha FW, Protopopoff N, Messenger LA., 2021. An increasing role of pyrethroid-resistant Anopheles funestus in malaria transmission in the Lake Zone, Tanzania. Sci Rep 11: 13457

33. Rehman AM, Coleman M, Schwabe C, Baltazar G, Matias A, Gomes IR, Yellott L, Aragon C, Nchama GN, Mzilahowa T, Rowland M, Kleinschmidt I., 2011. How much does malaria vector control quality matter: the epidemiological impact of holed nets and inadequate indoor residual spraying. PloS One 6: e19205

34. Mogeni P, Williams TN, Fegan G, Nyundo C, Bauni E, Mwai K, Omedo I, Njuguna P, Newton CR, Osier F, Berkley JA, Hammitt LL, Lowe B, Mwambingu G, Awuondo K, et al., 2016. Age, Spatial, and Temporal Variations in Hospital Admissions with Malaria in Kilifi County, Kenya: A 25-Year Longitudinal Observational Study. PLoS Med 13: e1002047

35. Bouyou-Akotet MK, Mawili-Mboumba DP, Kendjo E, Mabika-Mamfoumbi M, Ngoungou EB, Dzeing-Ella A, Pemba-Mihindou M, Ibinga E, Efame-Eya E, MCRU team, Planche T, Kremsner PG, Kombila M., 2009. Evidence of decline of malaria in the general hospital of Libreville, Gabon from 2000 to 2008. Malar J 8: 300

36. West PA, Protopopoff N, Rowland M, Cumming E, Rand A, Drakeley C, Wright A, Kivaju Z, Kirby MJ, Mosha FW, Kisinza W, Kleinschmidt I., 2013. Malaria Risk Factors in North West Tanzania: The Effect of Spraying, Nets and Wealth. PLOS ONE 8: e65787

37. Woolhouse ME., 1998. Patterns in parasite epidemiology: the peak shift. Parasitol Today Pers Ed 14: 428–434

38. Snow RW, Marsh K., 2002. The consequences of reducing transmission of Plasmodium falciparum in Africa. Adv Parasitol 52: 235–264

39. Nkumama IN, O’Meara WP, Osier FHA., 2017. Changes in Malaria Epidemiology in Africa and New Challenges for Elimination. Trends Parasitol 33: 128–140

40. Roca-Feltrer A, Carneiro I, Smith L, Schellenberg JRA, Greenwood B, Schellenberg D., 2010. The age patterns of severe malaria syndromes in sub-Saharan Africa across a range of transmission intensities and seasonality settings. Malar J 9: 282

41. Paton RS, Kamau A, Akech S, Agweyu A, Ogero M, Mwandawiro C, Mturi N, Mohammed S, Mpimbaza A, Kariuki S, Otieno NA, Nyawanda BO, Mohamed AF, Mtove G, Reyburn H, et al., 2021. Malaria infection and severe disease risks in Africa. Science 373: 926–931

42. Andolina C, Rek JC, Briggs J, Okoth J, Musiime A, Ramjith J, Teyssier N, Conrad M, Nankabirwa JI, Lanke K, Rodriguez-Barraquer I, Meerstein-Kessel L, Arinaitwe E, Olwoch P, Rosenthal PJ, et al., 2021. Sources of persistent malaria transmission in a setting with effective malaria control in eastern Uganda: a longitudinal, observational cohort study. Lancet Infect Dis 21: 1568–1578

43. Cohee LM, Nankabirwa JI, Greenwood B, Djimde A, Mathanga DP., 2021. Time for malaria control in school-age children. Lancet Child Adolesc Health 5: 537–538

44. Coalson JE, Cohee LM, Buchwald AG, Nyambalo A, Kubale J, Seydel KB, Mathanga D, Taylor TE, Laufer MK, Wilson ML., 2018. Simulation models predict that school-age children are responsible for most human-to-mosquito Plasmodium falciparum transmission in southern Malawi. Malar J 17: 147

45. Degarege A, Fennie K, Degarege D, Chennupati S, Madhivanan P., 2019. Improving socioeconomic status may reduce the burden of malaria in sub Saharan Africa: A systematic review and meta-analysis. PloS One 14: e0211205

46. Yang G, Kim D, Pham A, Paul CJ., 2018. A Meta-Regression Analysis of the Effectiveness of Mosquito Nets for Malaria Control: The Value of Long-Lasting Insecticide Nets. Int J Environ Res Public Health 15: 546

47. Ebenezer A, Noutcha AEM, Okiwelu SN., 2016. Relationship of annual entomological inoculation rates to malaria transmission indices, Bayelsa State, Nigeria. J Vector Borne Dis 53: 46–53

48. Smith DL, Dushoff J, Snow RW, Hay SI., 2005. The entomological inoculation rate and Plasmodium falciparum infection in African children. Nature 438: 492–495

49. Tokponnon FT, Ogouyémi AH, Sissinto Y, Sovi A, Gnanguenon V, Cornélie S, Adéothy AA, Ossè R, Wakpo A, Gbénou D, Oke M, Kinde-Gazard D, Kleinschmidt I, Akogbeto MC, Massougbodji A., 2014. Impact of long-lasting, insecticidal nets on anaemia and prevalence of Plasmodium falciparum among children under five years in areas with highly resistant malaria vectors. Malar J 13: 76

50. Protopopoff N, Rowland M., 2018. Accelerating the evidence for new classes of long-lasting insecticide-treated nets. Lancet Lond Engl 391: 2415–2416

51. Yahouédo GA, Cornelie S, Djègbè I, Ahlonsou J, Aboubakar S, Soares C, Akogbéto M, Corbel V., 2016. Dynamics of pyrethroid resistance in malaria vectors in southern Benin following a large scale implementation of vector control interventions. Parasit Vectors 9: 385

52. Sovi A, Govoétchan R, Ossé R, Koukpo CZ, Salako AS, Syme T, Anagonou R, Fongnikin A, Nwangwu UC, Oké-Agbo F, Tokponnon F, Padonou GG, Akogbeto MC., 2020. Resistance status of Anopheles gambiae s.l. to insecticides following the 2011 mass distribution campaign of long-lasting insecticidal nets (LLINs) in the Plateau Department, south-eastern Benin. Malar J 19: 26

53. Protopopoff N, Mosha JF, Lukole E, Charlwood JD, Wright A, Mwalimu CD, Manjurano A, Mosha FW, Kisinza W, Kleinschmidt I, Rowland M., 2018. Effectiveness of a long-lasting piperonyl butoxide-treated insecticidal net and indoor residual spray interventions, separately and together, against malaria transmitted by pyrethroid-resistant mosquitoes: a cluster, randomised controlled, two-by-two factorial design trial. The Lancet 391: 1577–1588

54. Staedke SG, Gonahasa S, Dorsey G, Kamya MR, Maiteki-Sebuguzi C, Lynd A, Katureebe A, Kyohere M, Mutungi P, Kigozi SP, Opigo J, Hemingway J, Donnelly MJ., 2020. Effect of long-lasting insecticidal nets with and without piperonyl butoxide on malaria indicators in Uganda (LLINEUP): a pragmatic, cluster-randomised trial embedded in a national LLIN distribution campaign. Lancet Lond Engl 395: 1292–1303

